# Data Driven Monitoring in Community Based Management of SAM children using Psychometric Techniques: An Operational Framework

**DOI:** 10.1101/2021.06.16.21258807

**Authors:** Ankur Joshi, Abhijit P Pakhare, Sivaja K Nair, G Revadi, Manoj Chouhan, Deepak Pandey, Arun M Kokane

**Affiliations:** Department of Community & Family Medicine AIIMS Bhopal; Ex CSAM Programme coordinator; MSW, Field monitor CSAM programme, AIIMS Bhopal; Regional Field monitor CSAM programme, AIIMS Bhopal

**Keywords:** Severe Acute Malnutrition, RASCH assessment, data driven monitoring

## Abstract

**Background:** The success of the Community Based Management of Severe Malnutrition (CSAM)programme, largely depends on the knowledge and skills of Front-Line Workers (FLWs).A robust supportive supervision system in CSAM should be tailored to individualistic learning needs by distinguishing the FLWs as per their ability and simultaneously identifying the task domains to be emphasized more in supervisory visits.This paper details the ability assessment strategy developed and employed in the selected geographical locations in state of Madhya Pradesh (Central India) among the 197 Anganwadi workers (FLWs involved in CSAM implementation)

**Methodology:** A 25 items tool was developed based on an analytical construct for ability estimation through Rasch Analysis (RA). RA models the probability of right/wrong answer as a function of person(participants) and item (questions) parameters and calculates the item difficulty in relation with person ability on same unidimensional linear scale. The fitting of the data to Rasch model (Rasch diagnostic) was tested by both numeric (Anderson LR and Wald test) and graphical method. Suitable visualization like Item Characteristic Curve (ICC) and Person Item Map (PIM) were plotted in RA.

Further a quadratic allocation of all AWWs into 4 quadrants were done as per the ability estimation (Rasch score) and adjusted numbers of SAM/MAM children in her center.

**Results:** The item easiness parameter (β) value related to Diarrhoeal assessment was lowest (−2.32, -2.91 to -1.73) and related to peer assessment consequential action (2.009, 1.669-2.348)) was highest (most difficult). Anderson LR test (LR=31.32, df=24, p=0.079) showed the absence of global outliers. Quadrant analysis using the permutations of ability score and adjusted burden of malnutrition further mapped 41/197 (20.8%) FLWs to low ability -high burden quadrant and 44/197(25%) as low ability low burden quadrant.

**Conclusion:** RASCH assessment may address the innate challenges to maintain homogeneity, discrimination capacity and linearity in a raw score-based measurement construct. The monitoring strategy developed on this thus may offer a judicious, pragmatic and thematic approach to supportive supervision in CSAM program.

## Introduction

Severely Acute Malnutrition (SAM) is a condition characterized by extreme low weight for Height, muscles wasting and nutritional oedema and is closely associated with high rates of mortality and morbidity among children under the age of five ^1^. A child with SAM or MAM (Moderate Acute Malnutrition) has more likelihood of mortality than well-nourished child if not intervened in a timely manner^2^.Till the first decade of this century, all such children were largely taken care of by a specifically designed intervention centers (Nutritional Rehabilitation Center -NRCs) having an assortment of medical, nutritional and educational interventions (FSAM strategy). However, this strategy was facing several challenges such as high operational and opportunity costs (for both systems and parents of SAM children), low coverage, longer stays leading to overcrowded NRCs, and cross-contamination^3,4,5^. Moreover, multiple studies have stated that only 10-20% of the SAM children had developed complications which required hospitalized care, whereas the rest could be treated at the community level with a package of services (medicines, nutritional supplements, nutrient dense foods and weekly tracking) ^3,6^. Taking these into account, an alternative solution to treat medically uncomplicated SAM children at their vicinity gradually emerged and Community Based Management of SAM children (CSAM) came into existence^7,8,9^. It is also known as Community based Management of Acute Malnutrition (CMAM) globally.

CSAM offers an opportunity for longitudinal tracking of the child by an intersectoral peripheral designated team at their doorsteps for a continuous monitoring of their nutritional status. Though this strategy was capable of addressing many challenges faced, many newer issues emerged which demanded immediate attention. A number of processes beginning from initial screening, classification, management and subsequent follow ups of the beneficiaries in CSAM are mostly performed by the peripheral Front-Line Worker (FLW) team constituted chiefly by Anganwadi Worker (AWW) and Auxiliary Nurse Midwife (ANM). This also includes the initial and follow up assessment for medical complications and subsequent referral to the health facility. Thus, the effectiveness of the program in principle depends on a robust ability of these FLWs to understand, apply and make decisions as per programmatic guidelines.

Thus, the monitoring and supervision strategies in this program must be sensitive enough to capture the extent of ability of FLWs about the conceptual understating and translating deliverables in CSAM strategy, without taking into consideration this aspect negates the principles on which they were built upon. The ability estimation is an abstract and latent construct in Psychometric paradigm. Generally, the measurement of this construct is attempted by some manifested variables (henceforth referred as items) which are presumed to measure the unique fragment of that construct. The consequential sum of these items is perceived as ability. This seemingly easy concept of ability measurement is little tricky on operational plane specially in social sciences /psychometrics. The raw scores derived by this summation may ignore the equidistance and linearity of items on difficulty level^13^. Thus these items may not discriminate the high from low achievers in true sense and neither offer an insight to evaluator whether he is measuring accurately a homogeneous construct only^14,15,16^There are multiple studies on the assessment of skills and capacities of the frontline workers; a few regarding programmes pertaining to management of malnutrition and few others on generic public health programmes^10,11,12^. However, majority of them estimate the ability by raw score without any transformation which neither takes into account the item difficulty and associated discrimination capacity of item nor it addresses the equidistance distribution of items and participants on a linear scale. Translating it from programmatic stance, the differential capacities of FLWs on different domains of tasks may not be explicitly addressed through them.

An alternate strategy to estimate the ability is offered by Rasch Analysis (RA) which takes into account the problem of homogeneity, discrimination capacity and linearity of items^17^. This is a mathematical modelling technique routed in logarithmic transformation. It attempts to achieve a conjoint (person -item measurement on the same linear scale) additivity (equi-distance linearity) while the homogeneity of the tool is simultaneously attempted to maximized by reduction of items (poor discriminator, misclassifies etc.)^18^. It presumes that probability of a correct response on an item is the product of calibrated item difficulty and calibrated person ability measured on logarithmic scale. The further description is of this scale is given in supplementary file.

Within the context above, we developed a data driven monitoring strategy which considered malnutrition magnitude with reference to ability (measured by RA) estimation. The objectives for this strategy development were to identify the AWCs requiring frequent supportive supervisory visits and to identify the programmatic components to be addressed during the visit.

## Methodology

This study was designed by CSAM unit of Regional Center of Excellence for Nutritional Rehabilitation and Resource Training (RCoENRRT), Madhya Pradesh. The RCoE is established under the aegis of AIIMS Bhopal, an apex teaching tertiary care hospital in Central India. The RCoE has adopted a sub-district development block (Babai) from the district of Hoshangabad for technical facilitation, execution and supportive supervision of CSAM program. The methodology section is further subdivided into three different subsections for ease of understanding.

### A1 Tool development

The process of tool development started in the month of September 2019. The first step undertaken in the process of tool development was to identify the theoretical construct from literature review and in-house discussions among investigators. This process led to the identification of three domains namely performance, adaptability and stability that could be further contextualized for CSAM management^19,20^. The initial desk review was followed by 4 field visits and engagement with the field experts for refining identifying the subdomains that would be specific to the context. The identified subdomain within the analytical framework is detailed in Table 1.

**Table 1:**
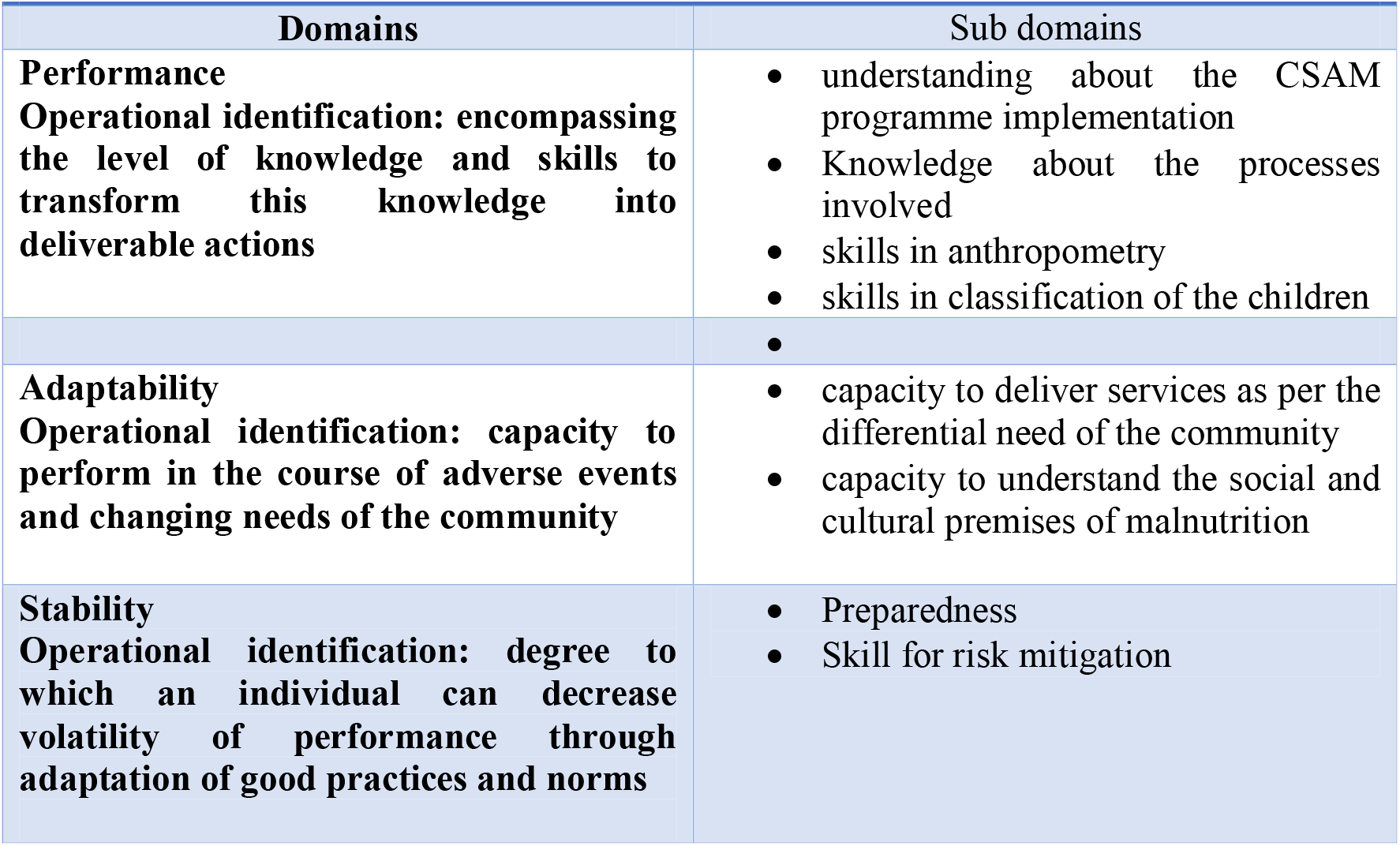
The domains and subdomains of assessment Analytical construct defining domains and subdomains for the study.

**Table 2.** Item difficulty parameters point estimates with 95% CI. The more the negative value the easier the item.

| Item Code | Item Description | Beta estimate | Lower CI | Upper CI |
| --- | --- | --- | --- | --- |
| V2 | Right Classification | 0.076 | -0.23 | 0.382 |
| V3 | Diarrhoea Detection - subsequent action | -2.319 | -2.906 | -1.732 |
| V4 | Length measurement sequence | 0.648 | 0.353 | 0.944 |
| V5 | Height measurement tool identification | -1.344 | -1.761 | -0.928 |
| V6 | Age identification method | -1.205 | -1.605 | -0.805 |
| V7 | Weight measurement tool identification | -1.659 | -2.12 | -1.197 |
| V8 | Weight measurement sequence | -0.103 | -0.416 | 0.21 |
| V9 | Previous h/o SAM in family -action | -1.344 | -1.761 | -0.928 |
| V10 | Target weight not achieve-action | 0.486 | 0.189 | 0.782 |
| V11 | Yellow cat classification-action | -0.965 | -1.338 | -0.591 |
| V12 | THR refusal -action | -0.292 | -0.614 | 0.03 |
| V13 | Oedema identification | 1.56 | 1.246 | 1.873 |
| V14 | Wrong statement-classification yellow/red | 1.234 | 0.932 | 1.537 |
| V15 | Migration of SAM child-action | -0.684 | -1.032 | -0.335 |
| V16 | Wrong statement -Appetite test | 0.95 | 0.653 | 1.247 |
| V17 | Medical complication in SAM | 0.051 | -0.256 | 0.358 |
| V18 | THR sharing family -action | -0.495 | -0.83 | -0.16 |
| V19 | Peer assessment scenario | 2.009 | 1.669 | 2.348 |
| V20 | Classification with weight /height | 0.344 | 0.045 | 0.643 |
| V21 | Right statement -THR quantity | 0.695 | 0.399 | 0.99 |
| V22 | Follow up length after discharge | 0.462 | 0.165 | 0.759 |
| V23 | Miscalculated Iron syrup dose | 0.672 | 0.376 | 0.967 |
| V24 | Difference-Wasting Stunting | 1.978 | 1.641 | 2.316 |
| V25 | Wrong statement -CSAM execution | 0.368 | 0.069 | 0.666 |

After identifying the subdomains, the research team decided to develop the items and transect the items as vital, essential and desirable matrix for each subdomain. We had developed 36 items initially based on the subdomains and the items were put to discussion with the research and field experts for deciding on the validity of the content, structure of the contents, and the length of the questionnaire. The items were proposed to be closed ended with multiple choices. This review of the initial pool of items, reduced the number of questions to 30, once the redundant and repetitive questions were deleted from the list and few of the questions were changed as per the expert opinion considering its contextual applicability. These questions were further translated into the local language with the help of a field expert proficient in the vernacular language. The translated items were put for pre-test with 7 research participants to check for issues in the language, tone, structure, and design of a questionnaire. Incorporating the suggestions from the research participants and the identified need, changes were made to the questions and the number of items were reduced to 27. A round of pilot test was undertaken to assess the consistency and reception of question and relevant changes were made to the pool of items and the item numbers were finalized as 25. Another round of pilot test was undertaken with the items before finalizing the tool to assess clarity and ease of understanding of the questions.

Information on burden of malnutrition for each Anganwadi centre was retrieved from monthly report submitted at block office. Number of children with Severe Acute Malnutrition (SAM-Weight for Height Z-score <-3) and Moderate Acute Malnutrition (MAM-Weight for Height Z-score <-2) were noted.

### A2 -Tool institution

The pretested and piloted tool was ready to use by the month of October 2019. It was a community based cross sectional psychometric health system research. As planned and communicated earlier, the Anganwadi workers (AWW) of Babai block were called for a block level review meeting on CSAM programme to the Block Panchayath, through the Child Development Project Officer (CDPO) on 11 October 2019. It was decided to conduct the RASCH assessment prior to the review meeting. Out of 217 AWWs in Babai block, 197 AWWs were present for the exercise. The pre-printed questionnaires were handed out to the AWWs. None of the questions were explained or read out to the research participants even when in doubt, since an individual level of understanding of the question was the mandate for the assessment. It was also communicated with the participants not to have any discussions on the questions. All possible measures including a time frame of 30 minutes for the completion of the questionnaire was set to avoid potential bias in the execution process.

### A3 Analysis and model creation

The data was entered in MS Excel in wide data format and was checked for duplication, missing values, outliers and redundancies. The adjusted total burden of malnutrition was calculated for each Anganwadi Centre (AWC-Geo-administrative unit for AWW) by assigning half of the weight (0.50) to a MAM child compared to a SAM child (1). The analysis comprises of 25 items and 197 Anganwadi Workers (AWWs). The analysis was performed in base-R and r-packages available in open domains. For each item difficulty parameters with confidence intervals were obtained. Item Characteristic Curves (ICCs) were drawn for each domain item which showed the plot between the probability of correct response for an item on y -axis and underlying ability for the domain-item. The purpose was to have a visual idea of discrimination capacity and difficulty of the items clubbed under the domain construct as mentioned above.

A person -item map was plotted through *eRM* package. This map displays the range and position of item (lower panel) and person parameters (upper panel) on the same axis of latent dimension.

The Rasch model was initially created for 25 items observations on 197 persons. The fitting of the data to Rasch model (Rasch diagnostic) was tested by both numeric (Anderson LR and Wald test) and graphical method. The reason behind choosing the two tests was to look at the model both from global (all items simultaneously) and in individualistic item vise perspective. The Anderson LR statistic checked for the item bias (Differential Item Functioning globally) while Wald test looked for the individual item invariance by dividing the sample into two groups. The visual diagnostic was performed by using Goodness of Fit (GOF) plot. Here underlying assumptions of homogeneity (with confidence bend) was visualized by comparing the beta scores of two groups identified by raw score≤ median and median≥ raw score. Rasch model was also scrutinized to check for deterministic patterns by infit MSQ value. All the diagnostic was run on eRM R-package.

A scatter diagram was drawn with *ggplot2* package in which each co-ordinate represented the adjusted burden of malnutrition in AWC at X-axis and RASCH value scored by corresponding AWW at Y -axis. The malnutrition burden in a centre was though off in terms of total weighted number of SAM and MAM children enrolled in centre at the time of tool administration. A MAM child was given half of the wight compared to SAM child for calculating burden which ensured the relative high contribution of SAM into total burden. This scatter plot was further divided into four quadrants by placing a horizontal and vertical line intersecting at y and x axis. These intersecting lines represented the median values of Rasch score (horizontal line) and adjusted burden of malnutrition (vertical line). This exercise led to formation of four quadrants-Q1 (right lower)-low ability high burden, Q2(left lower)-low ability, low burden, Q3(left upper)-high ability, high burden and Q4(right upper)-high ability, low burden.

The following diagram illustrates the process from construct identification to model creation in the sequential manner-

**Figure 1:**
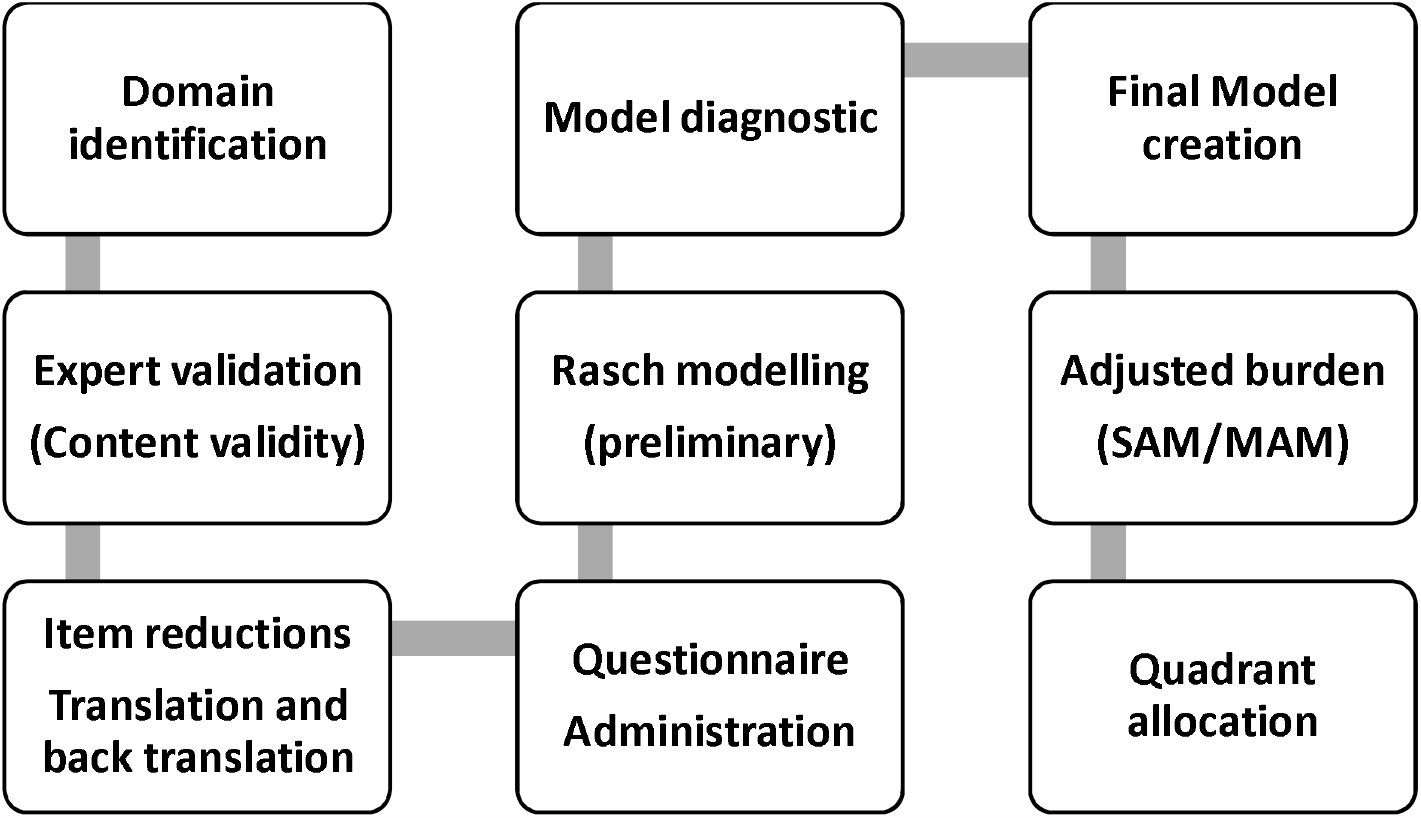
From Construct identification to model creation

### Ethics issues

Institutional Human Ethics Committee of AIIMS Bhopal reviewed and approved the study protocol. Committee also granted waiver of written informed consent. All participants were provided information sheet which had details about objectives and procedures. It was assured that performance in the assessment did not have any unfavourable effect on their regular working.

There are no competing interests. It is a funded by UNICEF, Madhya Pradesh.

## Results

The r codes and the detailed results are attached as appendix. The salient finding is shared here. Out of 217 AWWs in Babai block, 197 completed assessment. Average age of participants was 39.8 years and average duration of experience as AWW was 11.4 years. The item related to Diarrhoeal assessment was found to be as easiest (−2.32, -2.91 to -1.73). Item related to peer assessment consequential action (2.009, 1.669-2.348)) was most difficult on item difficulty parameters. The Item Characteristic Curve (ICC) is shown in fig 2 in which items are shown as per their domain membership with mean item difficulty scores. This figure shows the items related to ‘stability’ domains were perceived easy by AWWs and it had less discriminatory ability between low and high ability AWWs. On the other hand, items related to adaptability and performance had higher discriminatory capacity and were perceived as difficult by AWWs.

**Figure 2:**
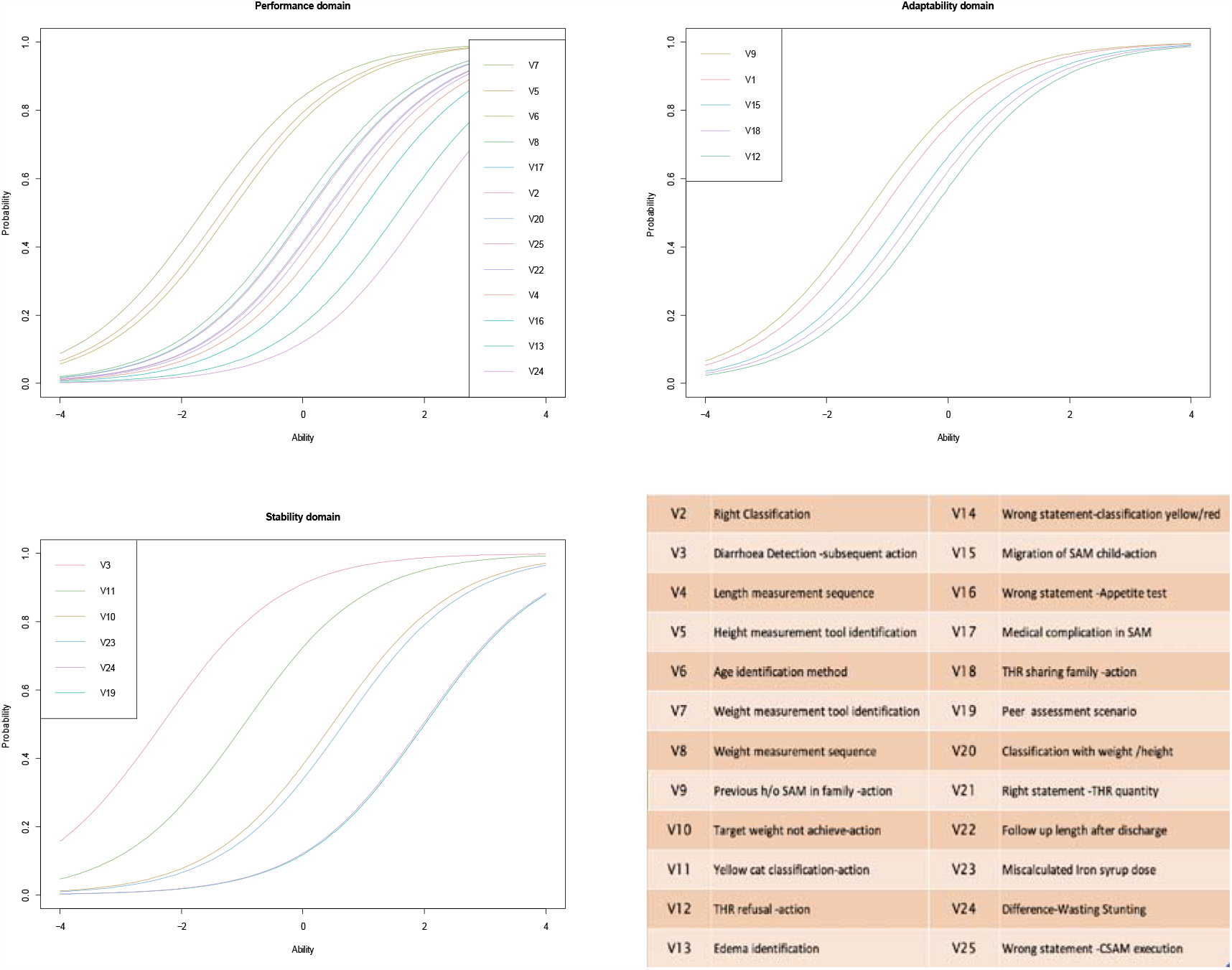
Item characteristic curve showing the monotonic homogeneity of ability on x -axis and probability to resolve at y axis across the 3 domains of inquiry

**Figure 3:**
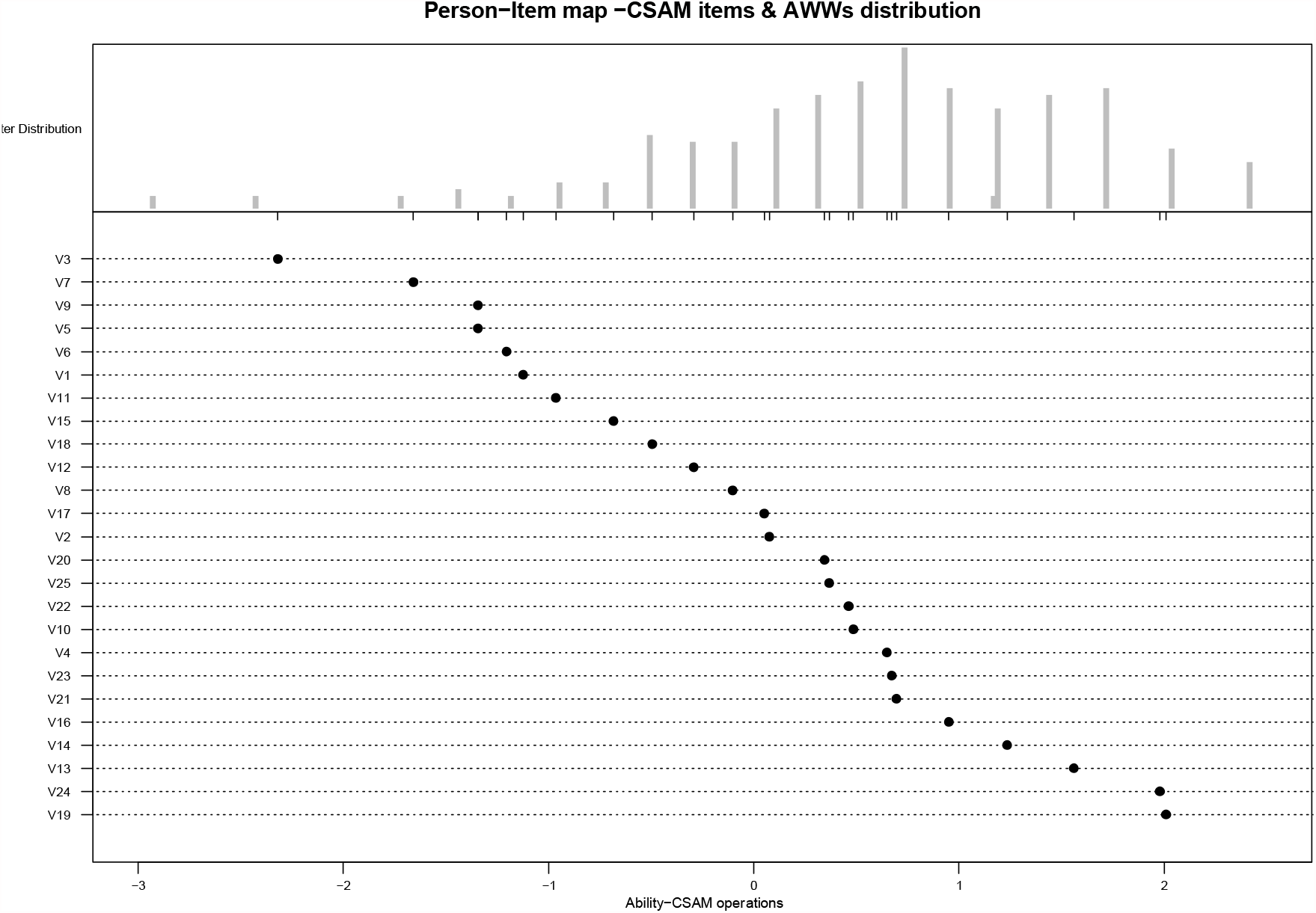
Person Item Map mapping the items and person along the same linear latent ability construct.

Person item map which maps the items and person on the same scale has shown that items were distributed along the whole range of latent dimensions thus sensitive enough to capture the ability of whole of the latent construct. However, this sensitivity was lower at very high ability (to the right upper end). Item no 4,21 and 23 mapped the maximum number of participants.

The purpose of the Rasch analysis is to assess the difficulty level apart from discriminatory capacity of items which is shown by ICC and Person Item map. The difficulty level can be assessed both at item and person levels both at same latent dimension. The next table shows the Item difficulty point estimates with their 95% confidence level.

Similarly, person parameters were also identified for 197 AWWs on the same latent dimension as of item parameter (individual person parameter values shown in supplementary file).

The descriptive analysis was followed by model diagnostic. Anderson LR test (LR=31.32, df=24, p=0.079) was applied to check the presence of global outliers which showed the absence of such outliers. Similarly, local outliers were checked through Wald statistic. 21 out of 25 items had p value more than the threshold while 4 items (item-12,19,24 and 25) had p values less than the threshold (<0.05). Yet these items were retained in the final model as they were in alignment with construct. Other tests/plots using in model diagnostics are shown in supplementary file.

The Rasch analysis and model diagnostic was followed by allocation of all the AWWs into 1 of the 4 quadrants created as under-low ability -high burden (P1-41 AWWs),low ability-low burden(P2-44 AWWs), high ability-high burden (P3-37AWWSs) and high ability -low burden(P4-75 AWWSs).This allocation of AWW as per the ability in CSAM domains and adjusted burden of the Malnutrition(differential weightage of MAM and SAM child) is shown through a visualization (fig-4) in which all the AWWs were categorized into one of the 4 quadrants .About 38% AWWs were having the high ability and low burden (low priority for monitoring with an inherent assumption of correct reporting as a function of ability) .Yet about 1 in5 AWWs had low ability and reported high burden of SAM c(P1) and about 1in 4 AWWs had low ability and low reported burden of SAM (P2).These P1 and P2 segments were though off as a priority segments for supportive supervision .This conceptual priority assignment was operationalized in 2 ways-enhanced relative allocation of monitoring visits to P1 and P2 segments and identification of difficult items from poor performing domains (by looking at item easiness parameters) with individualistic /group based capacity building.

**Figure 4:**
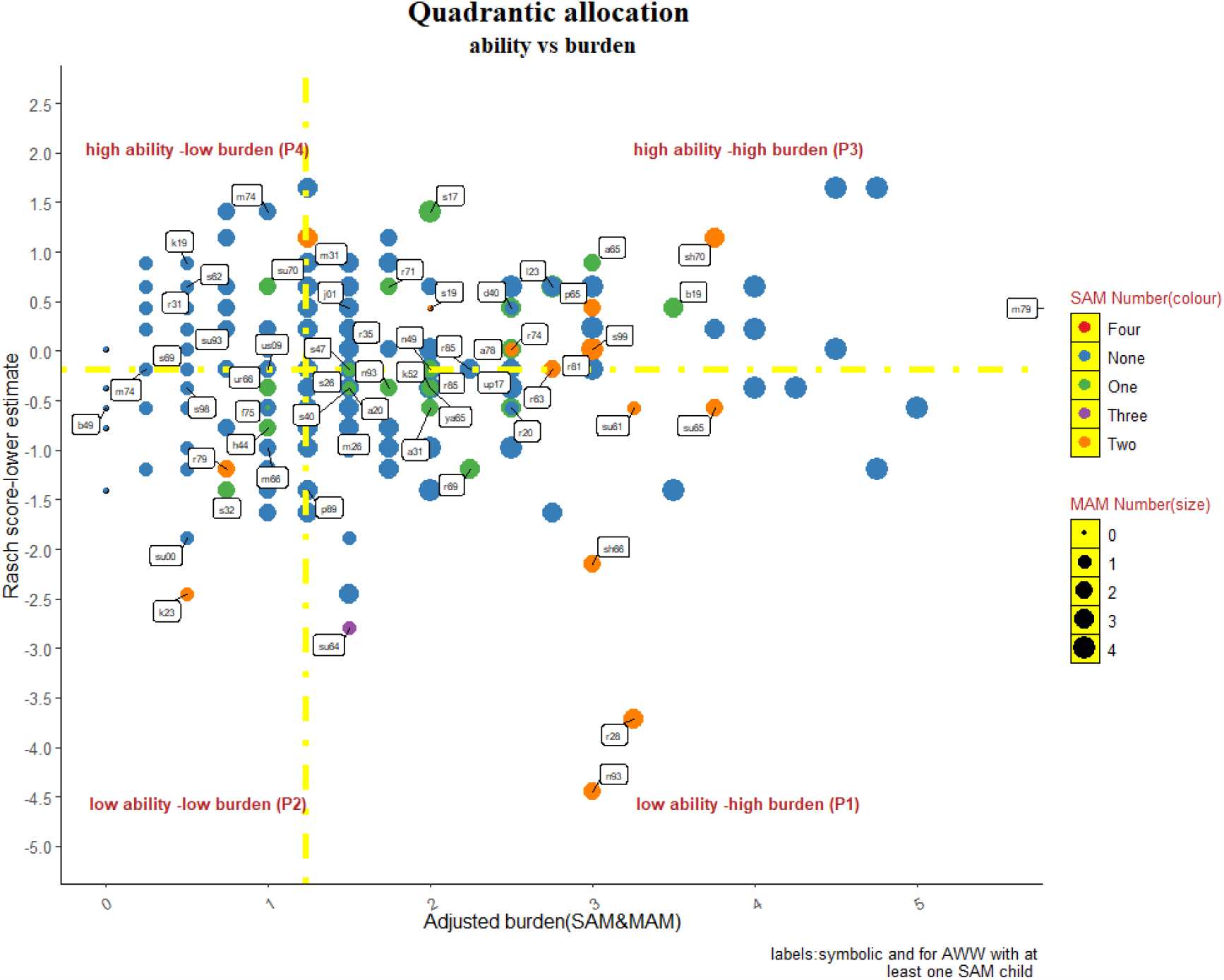
Quadratic allocation of AWWs as per ability (determined through RASCH) and adjusted burden of malnutrition. The yellow dashed lines intersect the x and y axis at their median value. The SAM number is colour coded while MAM numbers are shown by differential size of the dots.

**Figure 5:**
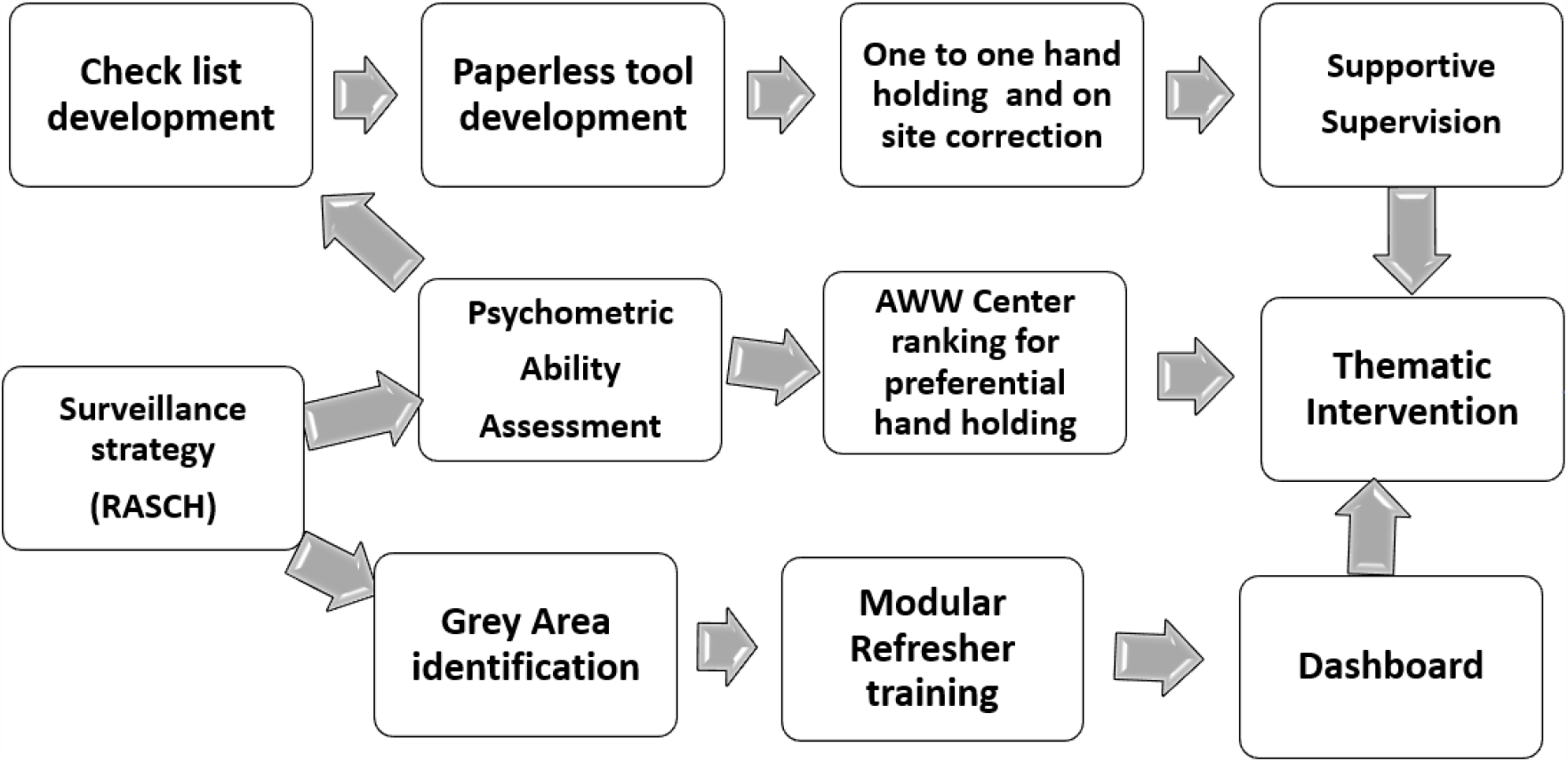
Intervention Framework

## Discussion

There has been a global shift in the way in which the process of supervision was perceived; from mechanistic authoritative supervision (“do as I say”) to supportive facilitatory (“do as we agree upon”) supervision^21,22^. To embark this paradigm, the supervisor should have the fair orientation of the strengths and scope of improvement of supervisee in their individualistic capacity and in an objective manner. In practice, it is often difficult to achieve due to previous cumulative predispositions and absence of tangible performance measures which have the factual and accurate construct of performance discrimination ^23,24^. Rasch modelling may attempt to overcome both concerns as it assigns an objectivity to whole evaluation framework and has an innate discriminant capacity for ability estimations. ^18^. In addition to that, any ability estimation in real time has to be translated into extent of task accomplishment by avoiding errors. With the argument, any monitoring strategy which simultaneously contemplates on ability and intended task may offer robust cues for actions compared to one which either focuses only on ability or intended task ^23,24,25^.

Aptness of any measurement depends on the notion of single construct (without contamination) equidistance and linearity in measurement in general. This is rather easy to achieve in physical measurements by concatenation. In the world of social science/psychometrics this measurement is allocation of numeral to events as per some pre-defined rubrics and the accumulation of these numbers is perceived as magnitude for that event ^26^. This simpler and linear calculation of the magnitude of an event will turn out to be problematic in social sciences since the observations do not fall in equidistant from each other and the positioning of a score has deeper meanings than its numerical value. A linear method of assessment might hence offer a flawed interpretation of the results and the understanding of the events in place ^18^. It is in this context that we have employed RASCH assessment as a psychometric tool to assess the ability of the Anganwadi workers in relation to the difficulty of the question in place.

A number of supervisory insights may be derived through this strategy-At first, in high burden and low ability quadrant, we will get to know whether this burden is a true burden or is it a anthropometric classification error on the part of field worker? This question is more relevant where there is a high discordance in MAM and SAM numbers. Although the discordance between SAM and MAM is reported by other population-based surveys and plausible explanations for the same is offered yet the transition from MAM to SAM should be perceived from the CAS (Complex Adaptive System) perspective where system outputs are the non-linear sums of underlying processes. and this transition should be seen on continuous patho-physiological spectrum from normal to SAM child and not as disconnected dichotomized entity. In the Quadrant plot the absolute SAM burden corresponds to colour of the marker thus adds to another dimension in the plot. Second, the low burden and low ability quadrant also conveys the same notion as mentioned above; whether are we missing some eligible SAM children here? The peripheral staff serves as ‘gate-keeper’ of CSAM management ecosystem through identification of SAM/MAM children. Thus, the delayed entry or missing cases may affect adversely the program performance thereby leading to preventable child mortality. Third, the high ability quadrants also have the high and low burden of the problem. These quadrants may serve as benchmarks for estimating the true burden but with some caveats. The cut-offs are derived from the median scores thus these quadrants are the outcome of rank order arrangement in two dimensions.

The standard error for Rasch score is maximum at the extreme ends and minimum at the middle which is opposite to raw scores. Ability estimation accordingly may have more reliability at middle compared to extreme ends ^27^. This argument although assigns more credibility to allocate the ‘grey -zone’ markers into respective quadrants yet farthest points at the quadrant plots should be interpreted with some caution. This inherent limitation of Rasch transformation may not affect substantially in our case as at person -item map, the item density is more at middle and non at the extreme end. Finally, the difficulty scores having agreements between predicted difficulties by investigators and actual difficulties are the definite domains in which capacity building is required. The RCoE has organized the refresher trainings for AWWs targeting specifically these domains both as groups and on one-to-one basis during the time of monitoring visits.

In summary, this strategy might offer some distinct advantages compared to raw scored based ability estimation as discussed above. The investigators have crafted the following intervention framework based on insights received from this data driven monitoring mechanism. However, the effectiveness of this framework will be achieved following the realistic application of focused monitoring and the improvement of the target indicators.

## Implications for policy and practice

We were able to identify the domain requiring more attention as well as we could develop a more focused monitoring strategy catering to the differential needs of the front-line workers. We further propose on the basis of this experience to explore the psychometric analysis tools (such as Rasch scores) for identification of specific activities to be focused for an individual as per his or her ability.

It also emphasizes consideration of burden of given health problem in deciding monitoring and supervision plan.

Thus, a data-driven framework proposed in this study will be helpful in prioritizing monitoring and supervision of outreach sites of community-based management of acute malnutrition. This strategy will ensure mentoring in identified individual specific domains.

The same framework can be suitably modified and adapted for monitoring and supervision of other public health programs.

## Supporting information

Supplementary File 1: RASCH questionnaire (English)

Supplementary File 2: RASCH Data file

Supplementary File 3: RASCH supplementary

Supplementary File 4: STROBE checkist

## Data Availability

The data will be provided in the supplementary files of the manuscript.

---

**Supplementary File 1 :** RASCH questionnaire (English)

**Supplementary File 2:** RASCH Data file

**Supplementary File 3:** RASCH supplementary

**Supplementary File 4:** STROBE checklist

