## Supplementary File 1: RASCH questionnaire (English) for "Data Driven Monitoring in Community Based Management of SAM children using Psychometric Techniques: An Operational Framework"

**Exploring the capacity building areas employing RASCH analysis**

AWW Centre No:

Village:

Name of the AWW:

Contact Number:

No of SAM Children registered at the centre in the current month/previous month:

Date of the interview:

1. Who is at maximum probability of risk of falling under SAM category
a. **Laxmi, fourth girl child of a daily wage laborer**
b. Ravi , only male child of a school teacher
c. Savitri , the second girl child of a political leader
d. Mukesh, male child having fever since 2 days
2. A 10 month old female child with 77 cm height and 6 Kg weight is having diarrhea and is unable to drink or be breastfed. How will you respond?
a. Give THR to the child
b. Register the child under CSAM activity
c. **Refer the child to the NRC**
3. A 36 month old normal male child is having severe diarrhea. How will you respond?
a. **Give ORS to the child**
b. Register the child under CSAM activity
c. Refer the child to the NRC
d. This child can be classified under MAM.
4. Which is the following depicted instrument will be used to measure the height of a 6 month old child? (IMAGE ONLY)

| 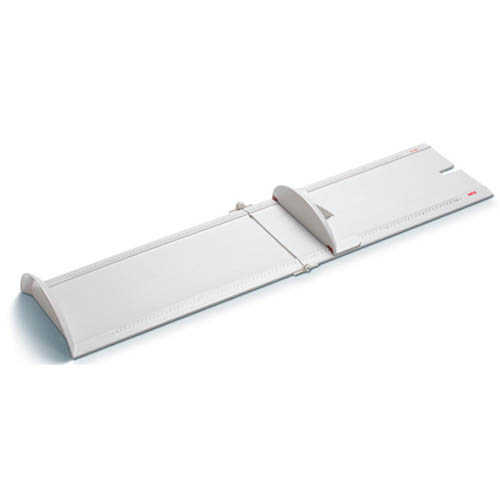 | 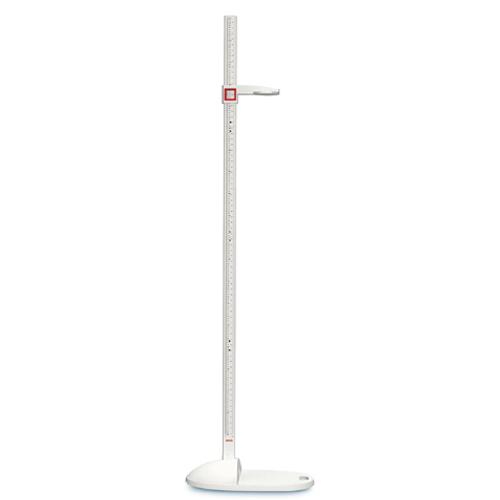 |
| --- | --- |
| 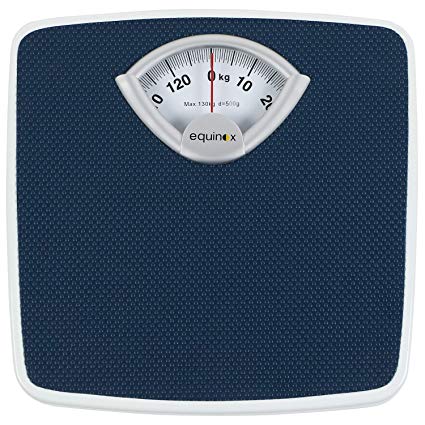 | 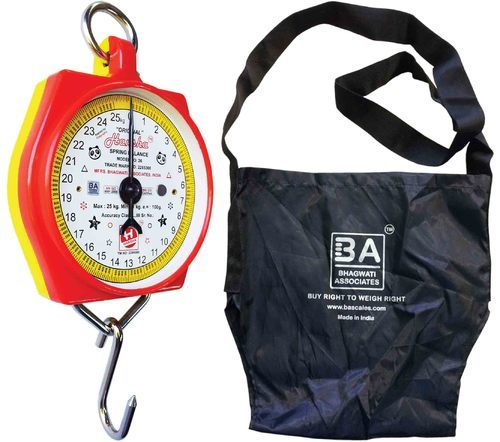 |

5. Which is the instrument used to measure the weight of a 15 month old child? (IMAGE only)

| 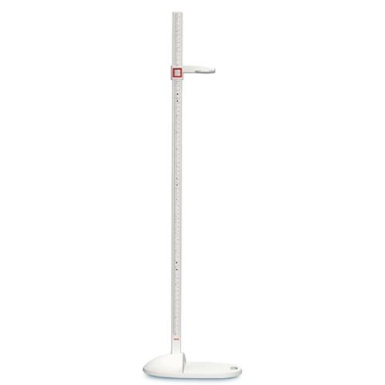 | 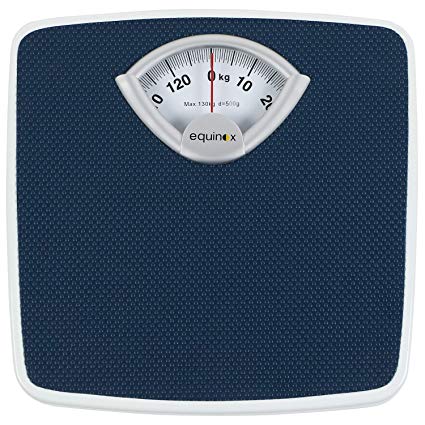 | 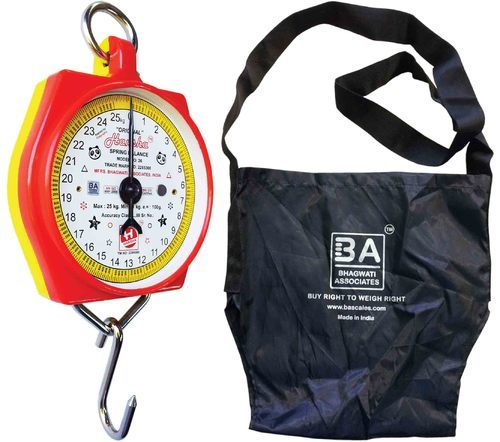 |
| --- | --- | --- |

6. Which is the correct order for measuring the height of a 26 month old child?

a. let the child lie down on the board on his/her back, position head against the headboard,

Knees and back straight, measure to the accuracy of 0.1 and note down the reading

b. **Remove their shoes, make the child stand against the stadiometer, Keep the knees together and**

**Straight, Keep the child’s head straight, Bring headpiece down onto the upper most point on the**

**child's Head, Record measurement to the last 0.1cm**

c. Remove their shoes, make the child stand against the stadiometer, Keep the child’s head straight, Bring

Headpiece down to the tip of the hair, Record measurement to the last 0.01cm.

7. If a new child has come to Anganwadi Center during CSAM session, how will you confirm the age of the

child.

a. By measuring the height of the child

b. By asking the mother

c. **By checking the birth certificate**

8. Which is the correct order for measuring the weight of an 18 month old child?

**a**. **Note the child’s age, Adjust the needle of the scale to zero, Put the child in the sling and hang**

**it on to the hook, Make sure the child’s feet are not touching the ground, Read the weight**

**when the child is calm and the needle stops moving**

**b**. Note the child’s age, Adjust the needle of the scale to zero, Put the child in the sling and hang

it on to the hook, Make sure the child’s feet are not touching the ground, hold on to the child

for safety, Read the weight when the child is calm and the needle stops moving

**c**. Remove heavier clothes on the child, Adjust the needle of the scale to zero and make the child stand on the digital scale and note down the measurement.

9. If a family has a history of SAM and the second girl child (1 year old) from the same family is presented as normal, what will be your response?

a. Refer the child to the NRC
b. Will make frequent visit to the family
c. Provide THR and monitor the feeding practices

10. If a child under the intervention hasn’t gained the desired weight in two months’ time, what will you do?

a. Follow up the child every week

b. Follow up the child every month

c. Provide more packets of THR

11. If the growth band of the child is in the yellow colour, what will you do?

a. Will register the child in the SAM category?

b. Will monitor the child and feeding practices

c. Will provide more packets of THR

12.Parents of an 18 month old female registered child having uncomplicated SAM has been denying to feed the child with THR, How will you respond to this?

a. Report to the supervisor

b. Will visit the child’s family, listen to their issues and try to educate them on the need

c. Register the child in the SAM category

d. Immediate intervene for correcting measure in a forceful way by calling the family to Aanganwadi

center

13. Which option is correct in the identification of bilateral pitting oedema

a. Press one feet of the child for 10 seconds and check for pitting

b. Apply thump pressure on the child’s feet for 3 seconds and check for pitting

c. Press both the feet of the child with both hands for 3 seconds and check for pitting

14. Tick which option is incorrect, if a child falls under the yellow or red zone in the growth chart

a. Inform the parents that their child's growth is slow for her/his age, Advise parents to pay greater attention on correct feeding practices and maintaining hygiene and cleanliness for their child.

b. Refer the child to the NRC

c. If the child is sick or weak, refer the child to the nearest NRC.

15. If a SAM child, was staying at her mother’s home, which is your duty station and have shifted her residence

to her father’s job location, How will you respond?

a. Report it to the supervisor

b. Report it to the NRC

c. Report it to the concerned anganwadi at the child’s current residence.

16. What is incorrect about conducting the appetite test?

a. The mother has to wash her hands before feeding the child

b. The child needs to be forced to complete the therapeutic food

c. It has to be made sure that the child was not fed for the last 2 hours.

17. Which of the following is a complicated SAM case

a. Child having Severe Dehydration

b. appetite pass

c. Child with edema in one foot

18. If you identify that the THR provided to a family is being shared among the whole family, how will you respond in this scenario (Tick the most appropriate)

a. Communicate to the parents regarding the need to feed THR to the child

b. Provide more packets of THR to the family as per family need

c. Will opt solely for monitored feeding of the child.

19. If a nearby AWW is unable to screen the child due to her incapacity and you came to know about this, how should you respond except-

a. Will communicate with the AWW and ask her to send the children to your centre for	screening

b. Will do nothing as any intervention may create confusion at administrative level.

c. Report it to the supervisor

d. will facilitate the help of ASHA of that village

20. Identify the child and categorize whether SAM, MAM or Normal.

11 month old male child with 60 cm height and 4.8 kg weight, No pitting edema but high fever

a. SAM
b. MAM
c. Normal

21. Which of the following is correct regarding the THR supply?

a. 5 Packet of THR per week for shalini who weighs 10.5 KG

b. 2 Packet of THR per week for Aman who weighs 7 kg

c. 3 Packet of THR per week for Raziya who weighs 9 Kg

22. What is the frequency of follow up visit after discharge from CSAM intervention?

a. Once very week

b. Once every month

c. Once in every fifteen days

23. If a child is under the CSAM intervention and you identify that the IFA and other medications are not given on correct dosage to the child. What will you do?

a. Teach the mother/ the caretaker about the correct dosage of the medicine

b. Supervised medication to ensure correct dosage

c. Report to supervisor

d All the above

24. Please tick on the statement which is right-

a. wasting is determined in reference to age.

b. Stunting is determined in reference to height.

c. Wasting denotes the present/ short time nutrition status

d. Stunting denotes the present/ short time nutrition status.

25. Which of the following is not true for CSAM program.

a. The target group for CSAM program is 6 -59 months children

b. CSAM session (satra) will be run at sub-center.

c. Registration will be done in CSAM center on VHSND in presence of ANM.

d. The risk of death by diseases like diarrhea is more in SAM child compared to normal child

26. Home visit by AWW is intended to check the compliance for following except-

a. THR diet

b. Amoxicillin and other drugs

c. Edema checking
