## Supplementary File 3: RASCH supplementary for "Data Driven Monitoring in Community Based Management of SAM children using Psychometric Techniques: An Operational Framework"

### RASCH Analysis

RCoE,CFM

1/25/2021

#### Loading of package and data

The datasheet consists of name of AWC,adjusted burden of SAM and MAM and response of 25 items(dicotomized )from 197 AWW.The RASCH analysis involes the *erm* package apart from base-r.

```
## Loading required package: MASS

## Loading required package: msm

## Loading required package: polycor

##
## Attaching package: 'dplyr'

## The following object is masked from 'package:MASS':
##
##     select

## The following objects are masked from 'package:stats':
##
##     filter, lag

## The following objects are masked from 'package:base':
##
##     intersect, setdiff, setequal, union
```

#### Rasch Priliminary modelling by using eRM package

##### Item parameters for both eta(difficulty) and beta (easiness)

```
##
## Results of RM estimation:
##
## Call:  RM(X = df1)
##
## Conditional log-likelihood: -2120.778
## Number of iterations: 25
## Number of parameters: 24
##
## Item (Category) Difficulty Parameters (eta): with 0.95 CI:
##     Estimate Std. Error lower CI upper CI
## v2     0.076      0.156   -0.230    0.382
```

```
## v3    -2.319      0.300   -2.906   -1.732
## v4     0.648      0.151    0.353    0.944
## v5    -1.344      0.212   -1.761   -0.928
## v6    -1.205      0.204   -1.605   -0.805
## v7    -1.659      0.235   -2.120   -1.197
## v8    -0.103      0.160   -0.416    0.210
## v9    -1.344      0.212   -1.761   -0.928
## v10    0.486      0.151    0.189    0.782
## v11   -0.965      0.191   -1.338   -0.591
## v12   -0.292      0.164   -0.614    0.030
## v13    1.560      0.160    1.246    1.873
## v14    1.234      0.154    0.932    1.537
## v15   -0.684      0.178   -1.032   -0.335
## v16    0.950      0.151    0.653    1.247
## v17    0.051      0.157   -0.256    0.358
## v18   -0.495      0.171   -0.830   -0.160
## v19    2.009      0.173    1.669    2.348
## v20    0.344      0.153    0.045    0.643
## v21    0.695      0.151    0.399    0.990
## v22    0.462      0.152    0.165    0.759
## v23    0.672      0.151    0.376    0.967
## v24    1.978      0.172    1.641    2.316
## v25    0.368      0.152    0.069    0.666
##
## Item Easiness Parameters (beta) with 0.95 CI:
##          Estimate Std. Error lower CI upper CI
## beta v1     1.123      0.199    0.733    1.513
## beta v2    -0.076      0.156   -0.382    0.230
## beta v3     2.319      0.300    1.732    2.906
## beta v4    -0.648      0.151   -0.944   -0.353
## beta v5     1.344      0.212    0.928    1.761
## beta v6     1.205      0.204    0.805    1.605
## beta v7     1.659      0.235    1.197    2.120
## beta v8     0.103      0.160   -0.210    0.416
## beta v9     1.344      0.212    0.928    1.761
## beta v10   -0.486      0.151   -0.782   -0.189
## beta v11    0.965      0.191    0.591    1.338
## beta v12    0.292      0.164   -0.030    0.614
## beta v13   -1.560      0.160   -1.873   -1.246
## beta v14   -1.234      0.154   -1.537   -0.932
## beta v15    0.684      0.178    0.335    1.032
## beta v16   -0.950      0.151   -1.247   -0.653
## beta v17   -0.051      0.157   -0.358    0.256
## beta v18    0.495      0.171    0.160    0.830
## beta v19   -2.009      0.173   -2.348   -1.669
## beta v20   -0.344      0.153   -0.643   -0.045
## beta v21   -0.695      0.151   -0.990   -0.399
## beta v22   -0.462      0.152   -0.759   -0.165
## beta v23   -0.672      0.151   -0.967   -0.376
```

```
## beta v24    -1.978       0.172    -2.316    -1.641
## beta v25    -0.368       0.152    -0.666    -0.069

## [1] -1.123091

##  beta v3  beta v7  beta v5  beta v9  beta v6  beta v1 beta v11 beta v15
##    -2.32    -1.66    -1.34    -1.34    -1.20    -1.12    -0.96    -0.68
## beta v18 beta v12  beta v8 beta v17  beta v2 beta v20 beta v25 beta v22
##    -0.49    -0.29    -0.10     0.05     0.08     0.34     0.37     0.46
## beta v10  beta v4 beta v23 beta v21 beta v16 beta v14 beta v13 beta v24
##     0.49     0.65     0.67     0.69     0.95     1.23     1.56     1.98
## beta v19
##     2.01
```

#### person-item map

A person-item map displays the location of item (and threshold) parameters as well as the distribution of person parameters along the latent dimension. Person-item maps are useful to compare the range and position of the item measure distribution (lower panel) to the range and position of the person measure distribution (upper panel).Items should ideally be located along the whole scale to meaningfully measure the ‘ability’ of all persons.

```
plotPImap(res_rm_1, item.subset = "all", sorted = TRUE,
          main = "Location of CSAM items distributions to AWWs distribution
along the latent constuct of ability",
          latdim = "CSAM Operational ability",
          pplabel = "AWWsPerson Paramter Distribution", cex.gen = 1,
          xrange = c(-3,2.5), warn.ord = TRUE, warn.ord.colour = "blue",
          irug = TRUE, pp = NULL)->pi_map
```

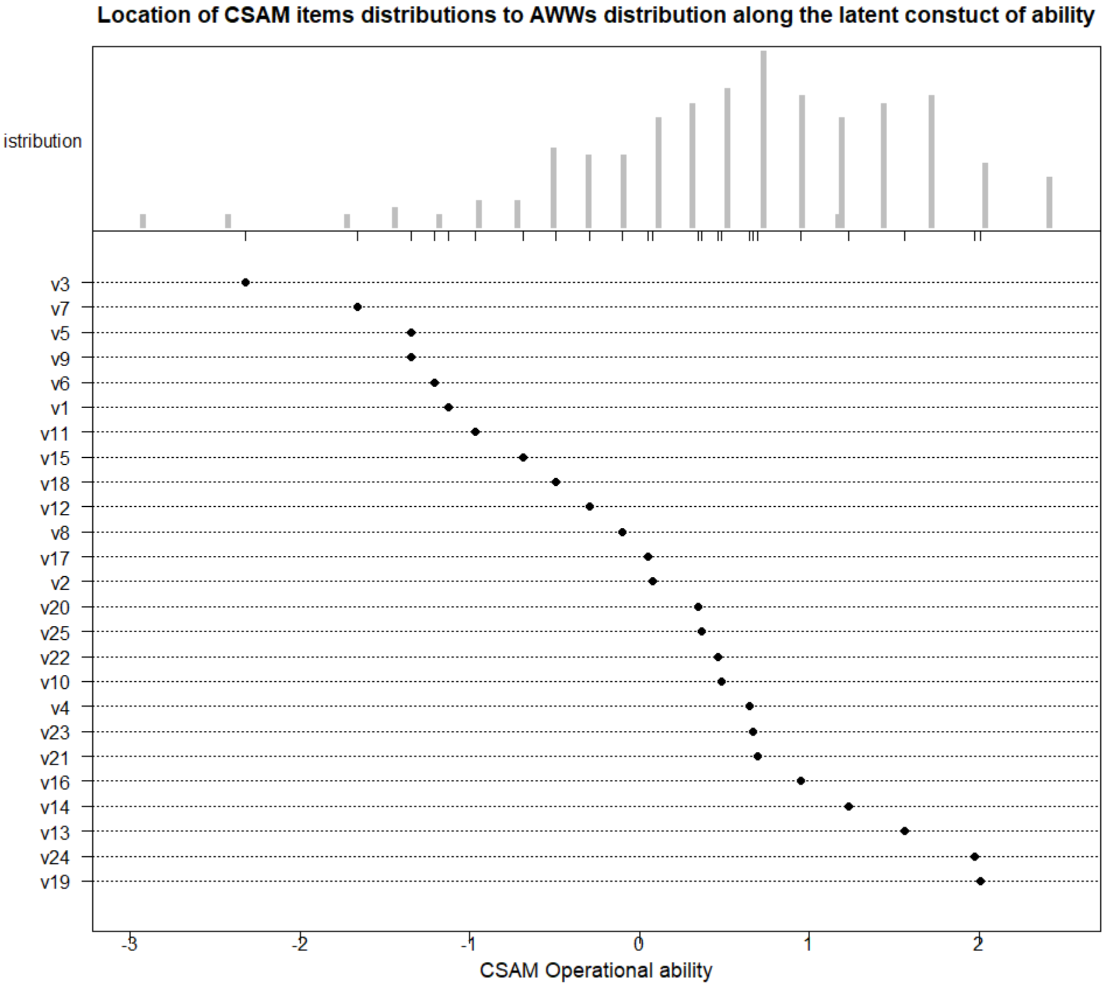


```
##Model Identificationidentification constraint,
##the identification constraint in RM() is a sum-to-zero constraint
round(sum(betas), 10)

## [1] 0
```

#### Rasch diagnostics

#### Andersen's Likelihood ratio test

```
test_data<-df1
test_data<-df1
model <- RM(test_data)
LR_median <- LRtest(model, splitcr = "median")
LR_median

##
## Andersen LR-test:
## LR-value: 34.315
```

```
## Chi-square df: 24
## p-value:  0.079
```

#Andersen LR-test:

### LR-value: 34.315 #Chi-square df: 24 #p-value: 0.079

####wald test chek for single items wt<-Waldtest(model) wt #q19, 24 and 25 have the large z value #after looking the construc there seems to be some typographical errors, in option b of q19 # q24 goes well with construct and will be included in final analyis #Question 25 mismatch with the construct and ‘satra’ and ‘up swathya kendra’ words needed to be look for.

### #those with large z-values in the Wald test . Do those items have special features that set them apart from the others in favoring one split group over the other?
